# Measuring community building in group antenatal care: Evidence from a randomized trial in Malawi

**DOI:** 10.64898/2026.11.10.26367565

**Authors:** Anne Batchelder, Ginger C Hanson, Esnath Kapito, Ellen Chirwa, Crystal L. Patil, Nancy Glass

## Abstract

Improving the quality of antenatal care is essential for reducing persistently high maternal and neonatal mortality in Malawi. Group antenatal care has demonstrated improvements in quality, satisfaction, health behaviors, and outcomes. However, the impact of community building, one of the model’s core components, has not previously been quantitatively assessed. This study examined the measurement properties of two community building measures, assessed the impact of group antenatal care on these measures, and explored their potential role as mediators of health outcomes.

This secondary analysis used quantitative data from the parent study testing the effectiveness of group antenatal care at seven clinics in Blantyre District, Malawi. The Perinatal Peer Support Scale and the Provider Connectedness subscale of the Pregnancy Related Empowerment Scale were used to measure community building and were validated using factor analyses. Results supported a two-factor structure for perinatal peer support (relational support and peer network) and a unidimensional structure for provider connectedness. Using generalized linear modeling, we found that participation in group antenatal care was associated with significantly stronger community building across all measures when compared to usual individual antenatal care. Women in group antenatal care demonstrated higher relational support score, peer network, and provider connectedness scores compared with women in individual antenatal care (*p* < 0.001). However, we found no evidence that community building had direct or indirect (mediating) effects on maternal psychological distress or infant birthweight.

This study strengthens the evidence base for group antenatal care by validating measures of community building and showing that relational support, peer networks, and provider connectedness are important benefits of participation. Although community building did not mediate the outcomes examined here, the findings highlight the need to further test of theoretical pathways linking group antenatal care to maternal and child health outcomes.

## Introduction

Malawi experiences some of the poorest maternal and infant outcomes in the world, with a maternal mortality rate of 439 women per 100,000 live births and an infant mortality rate of 46 infants per 1000 live births each year [1]. Improvements in the quality of antenatal care are needed, as low-quality antenatal care can contribute to higher rates of low-birth-weight infants, maternal and neonatal mortality, and poor mental health outcomes [2–7].

In Malawi, about 95% of pregnant women access antenatal care, which provides a point of contact to the health system; but only half of women attend all recommended visits [1,8]. Furthermore, women seeking antenatal care reported low-quality interactions with healthcare workers and high costs [9–11]. Additionally, antenatal care often requires long waits and short visits with the midwife, and women do not always experience respectful maternity care [2,3,12–14]. The Government of Malawi identified challenges to reducing maternal and infant mortality, including shortages in health funding, trained healthcare workers, and medical supplies [15].

CenteringPregnancy is an evidence-based model of group antenatal care that has the potential to reduce some of the challenges in antenatal care that contribute to maternal and infant mortality and morbidity in resource-limited settings. In 2016, Group antenatal care was recommended for research by the World Health Organization, and evidence from recent clinical trials in multiple African countries has found that group care models improve key outcomes, including facility birth and birth planning, antenatal care attendance, postpartum care engagement, quality of care, preparations for birth and awareness of potential complications, family planning knowledge, breastfeeding, health literacy, and mental health [16–22].

In Malawi, a CenteringPregnancy-based group antenatal care was adapted and implemented at seven clinics in Blantyre District. Pregnant women randomized to the group antenatal care intervention received antenatal care in a group setting facilitated by a trained Malawian midwife and community facilitators. Women randomized to individual antenatal care (usual care) were offered the eight antenatal care contacts recommended by the WHO on a first-come, first-served basis. While waiting together in the waiting room, they received a health lecture from a midwife. Then, they met individually with a midwife for a physical assessment. The full protocol and results from the effectiveness trial are available elsewhere [23,24].

CenteringPregnancy has three core components: healthcare in a group space, interactive learning, and community building, which are embedded in each of the sessions [25,26]. Assessments include individual health checks, where women measure and record their own weight and blood pressure and then meet with the midwife to review results (healthcare in group space), facilitated, educational conversations on topics including warning signs, birth preparation, partner communication, and nutrition (interactive learning), and socializing, interaction, and shared experiences with other pregnant women and providers (community building).

Bringing people together in group antenatal care and building peer and provider connections, could transform healthcare and outcomes for women, supported by evidence that social relationships affect health outcomes [27–31]. It is achieved through a stable group and time to socialize, which create trust, respect, safety, and belonging. These, in turn, allow women to be open and share information and experiences, and they grow into peer support and connection with providers [25]. Relationship-centered care is an important part of group antenatal care, and community building is how the relationships among the women and the women and the providers flourish, becoming a source of peer support and provider connectedness [25]. Grenier and colleagues’ theory of change frames group antenatal care as a transformed model of care that fosters a self-reinforcing cycle of increased, higher-quality care, improving health behaviors and maternal and newborn outcomes [32].

Group antenatal care has been shown to improve satisfaction and key outcomes, while also fostering trust and building relationships and community among participants. However, few studies have directly measured community building within group antenatal care. Understanding community building in group antenatal care may help to explain how and why group models work, especially in contexts like Malawi, where improving quality of antenatal care has been found to address high maternal and infant mortality and morbidity. Without understanding potential mechanisms contributing to effectiveness, such as community building, the scalability and adaptation of group antenatal care interventions may be limited by not understanding when interventions have high fidelity and what is creating improved results. This study addresses this gap by validating measures of perinatal peer support and provider connectedness to assess community building within a randomized controlled trial of group antenatal care in Malawi and to examine whether it mediates key maternal and infant outcomes.

The purpose of this study was to conduct a secondary analysis of data of the parent trial in Malawi. Specifically, the current study aimed to:

1. Measure community building in group antenatal care and its association with group assignment (intervention vs control)

a. Validate the Perinatal Peer Support Scale and Provider Connectedness subscale as a measure for community building
b. Examine the differences between group antenatal care and individual antenatal care on the measures of community building
2. Examine the mediating effect of community building on differences between group antenatal care and individual antenatal care on maternal (psychological distress) and infant (birth weight) outcomes

## Materials and Methods

### Study Design

This study is a secondary data analysis from a randomized controlled trial, “Group Antenatal Care: Effectiveness and Contextual Factors Linked to Implementation Success in Malawi” (Trial registration: ClinicalTrials.gov NCT03673709) to determine the effectiveness of a group antenatal care intervention on mothers and infants’ outcomes of birth weight and maternal mental health, compared to individual prenatal visits with midwife in Malawi. The protocol and results of the parent study are published elsewhere [23,24]. The data were first accessed on 06/19/2024.

### Setting

The parent study was implemented at seven antenatal care clinics in Blantyre District in Malawi, which is in the southern region of Malawi. Blantyre District is the home of the second largest city in Malawi and includes peri-urban and rural areas surrounding the city. The seven government-funded health centers provide health care to socioeconomically diverse populations from rural, peri-urban, and urban communities. The clinics also differed in the annual numbers of prenatal patients and midwives. Government funded clinics provide free health care to the populations they serve but also face substantial health workforce shortages, where only 40% of the national midwife positions are filled and there are only 1.48 health workers per 1,000 people, compared to the WHO’s minimum recommendation of 4.45 health workers per 1,000 people [33,34].

### Recruitment, Enrollment, and Consent

After a woman’s initial prenatal appointment, the midwives referred her to the parent study’s research assistants to learn about the study. If the women expressed interest, she was screened for eligibility. Pregnant women who were less than 24 weeks gestation, 15 years of age or older, and able to consent were eligible for the study. Women older than 17 provided written informed consent and women between the ages of 15-17 assented with written consent from a legal guardian as appropriate. Participants were consented to the study between July 5, 2019, and August 10, 2023. The secondary analysis was approved by Johns Hopkins Medical Institutions (IRB00355902). The parent study was approved by both the Kamuzu University of Health Sciences (COMREC: P.10/18/2498) and the University of Illinois Chicago (IRB: 2018–0845).

### Data Collection

Parent study data was collected longitudinally, at baseline during enrollment (T1) and at three follow up periods: at the end of the intervention (T2), two months postpartum (T3), and six months postpartum (T4). This secondary analysis included the entire sample (both intervention and control) and used data from T1 (covariates, intervention or control assignment), T2 (community building measures), and T3 (maternal and infant health outcomes).

### Measures

Table 1 shows all measures used in this study.

**Table 1:**
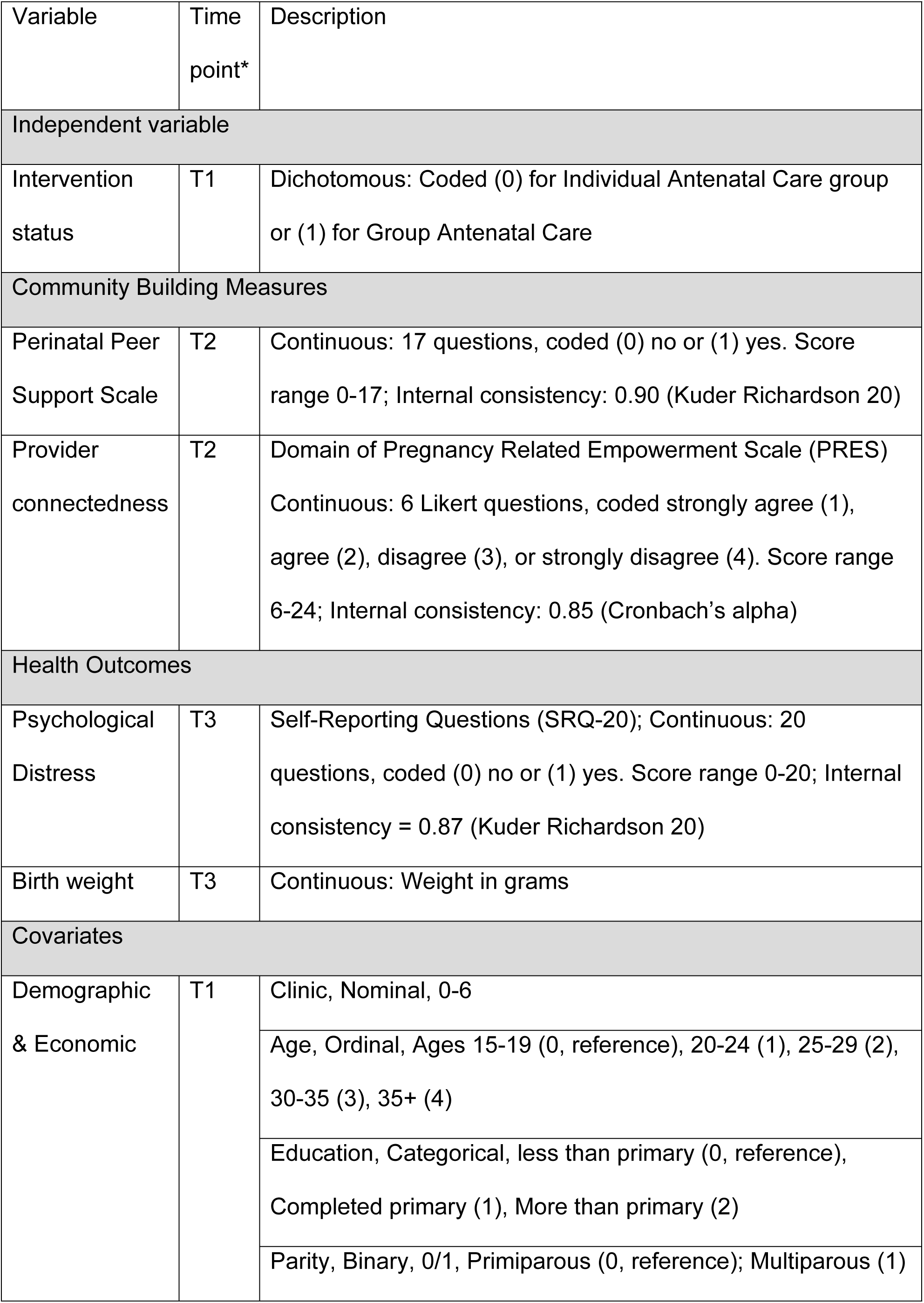
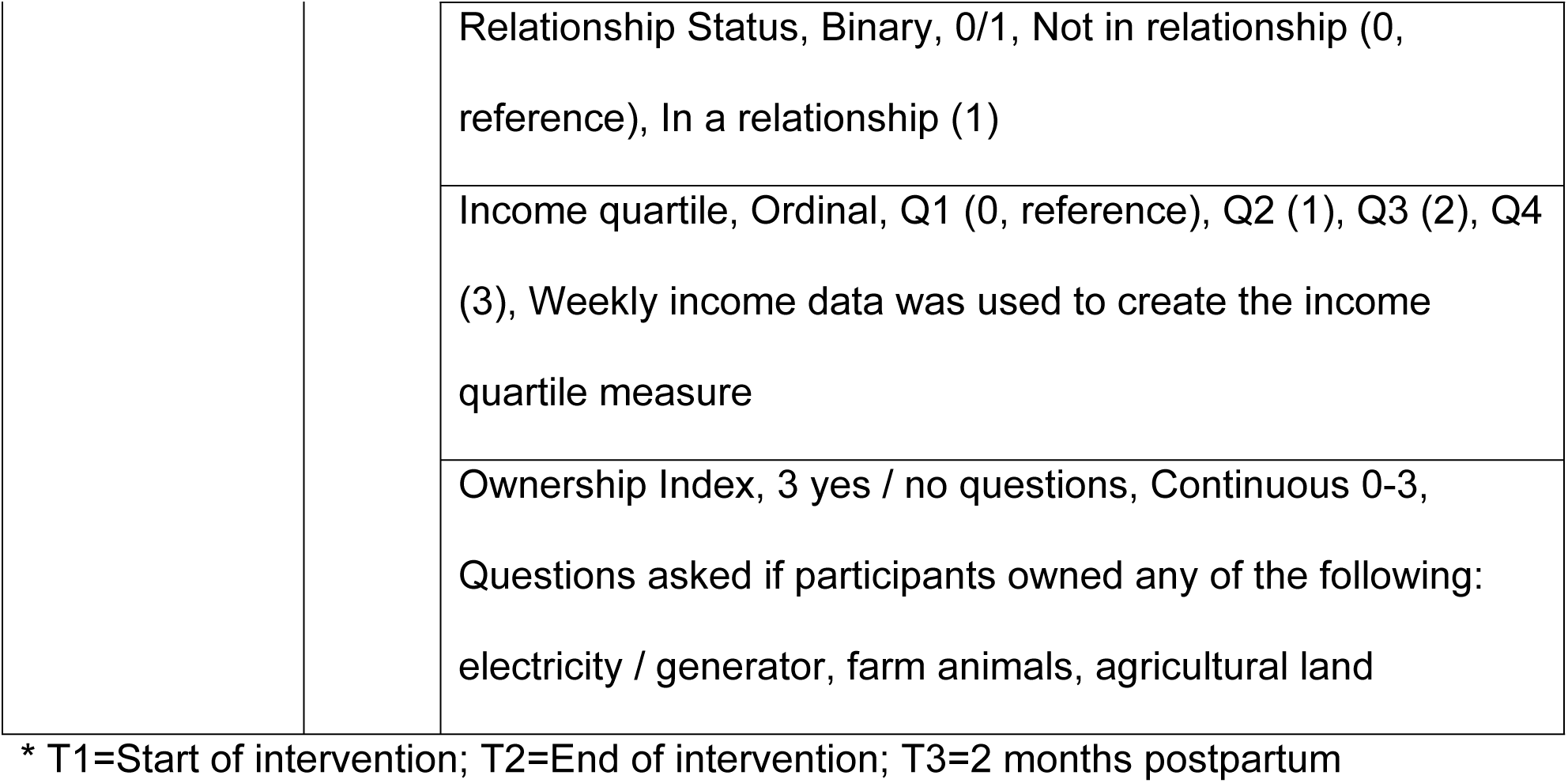
Measures.

| Variable | Time point* | Description |
|---|---|---|
| Independent variable |  |  |
| Intervention status | T1 | Dichotomous: Coded (0) for Individual Antenatal Care group or (1) for Group Antenatal Care |
| Community Building Measures |  |  |
| Perinatal Peer Support Scale | T2 | Continuous: 17 questions, coded (0) no or (1) yes. Score range 0-17; Internal consistency: 0.90 (Kuder Richardson 20) |
| Provider connectedness | T2 | Domain of Pregnancy Related Empowerment Scale (PRES)<br>Continuous: 6 Likert questions, coded strongly agree (1), agree (2), disagree (3), or strongly disagree (4). Score range 6-24; Internal consistency: 0.85 (Cronbach’s alpha) |
| Health Outcomes |  |  |
| Psychological Distress | T3 | Self-Reporting Questions (SRQ-20); Continuous: 20 questions, coded (0) no or (1) yes. Score range 0-20; Internal consistency = 0.87 (Kuder Richardson 20) |
| Birth weight | T3 | Continuous: Weight in grams |
| Covariates |  |  |
| Demographic & Economic | T1 | Clinic, Nominal, 0-6 |
|  |  | Age, Ordinal, Ages 15-19 (0, reference), 20-24 (1), 25-29 (2), 30-35 (3), 35+ (4) |
|  |  | Education, Categorical, less than primary (0, reference), Completed primary (1), More than primary (2) |
|  |  | Parity, Binary, 0/1, Primiparous (0, reference); Multiparous (1) |
|  |  | Relationship Status, Binary, 0/1, Not in relationship (0, reference), In a relationship (1) |
|  |  | Income quartile, Ordinal, Q1 (0, reference), Q2 (1), Q3 (2), Q4 (3), Weekly income data was used to create the income quartile measure |
|  |  | Ownership Index, 3 yes / no questions, Continuous 0-3, Questions asked if participants owned any of the following: electricity / generator, farm animals, agricultural land |

\* T1=Start of intervention; T2=End of intervention; T3=2 months postpartum

Consistent with this definition, community building was assessed with two measures reflecting its peer and provider dimensions: 1) perinatal peer support scale and 2) provider connectedness, a subscale of the Pregnancy Related Empowerment Scale (PRES). Both measures were developed for group-based care, while being worded to be applicable to those in individual care.

#### Perinatal Peer Support

During the pilot study predating the parent study, the group based antenatal care intervention was adapted to the Malawian context, and the perinatal peer support scale with 17 items was developed based on qualitative findings related to women’s experiences with peers in group care in order to measure how women in both individual and group antenatal care develop relationships with peers, experience emotional support, relate to each other, and exchange information [22]. The women randomized to individual antenatal care in the parent study were also receiving care at the seven participating clinics and likely had appointments on the same day and time as the women in group care, thus women may have shared learning through conversations as they waited for appointments and were often given a short educational program from midwives as a group during clinic visits. There is potential for women randomized to individual care from midwife to experience peer support. The individual items of the perinatal peer support scale are in Table 3.

**Table 2:** Participant demographic characteristics by individual antenatal care and group antenatal care groups.

| Characteristic, n (%) | Individual antenatal care (n = 941) | Group antenatal care (n = 946) |  |
|---|---|---|---|
|  | n (%) | n (%) | *p*-value |
| Primiparous (n = 1887) | 395 (42.1) | 379 (40.1) | 0.413 |
| Age (n=1884) |  |  | 0.617 |
| 15-19 | 261 (27.7) | 235 (24.8) |  |
| 20-24 | 353 (37.6) | 375 (39.7) |  |
| 25-29 | 190 (20.2) | 193 (20.4) |  |
| 30-34 | 87 (9.2) | 86 (9.1) |  |
| 35+ | 48 (5.1) | 56 (5.9) |  |
| In a relationships (n = 1887) | 874 (93.1) | 886 (93.9) | 0.491 |
| Education category (n = 1883) |  |  | 0.539 |
| Less than primary | 422 (44.8) | 414 (43.8) |  |
| Completed primary | 386 (41.0) | 381 (40.2) |  |
| More than primary | 133 (14.1) | 151 (16.0) |  |
| Ownership index (n = 1886)* |  |  | 0.815 |
| 0 | 191 (20.3) | 179 (18.9) |  |
| 1 | 454 (48.2) | 454 (48.0) |  |
| 2 | 254 (27.0) | 271 (28.7) |  |
| 3 | 42 (4.5) | 41 (4.3) |  |

\* Ownership index questions asked if participants owned any of the following: electricity/generator, farm animals, agricultural land.

**Table 3:** The Perinatal Peer Support Scale Items and Their Loading on Two Factors with Uniqueness.

|  | Factor 1: Relational Support | Factor 2: Peer Network | Uniqueness |
|---|---|---|---|
| 1) While at antenatal care, did you make new friends? | 0.13 | **0.68** | 0.41 |
| 2) Do you talk to any of these friends on the phone? | -0.16 | **0.86** | 0.40 |
| 3) Do you visit any of these friends? | -0.06 | **0.76** | 0.48 |
| *Instructions: As section will focus on relationships you developed with other women while attending clinic. First, you will hear a list of words that are about the kinds of emotional support that women sometimes get from one another. As you hear each word, please indicate if you experienced this type of support from other pregnant women at ANC.* |  |  |  |
| 4) Love/Affection | **0.55** | 0.36 | 0.32 |
| 5) Trust | **0.61** | 0.31 | 0.31 |
| 6) Acceptance | **0.61** | 0.25 | 0.37 |
| 7) Encouragement | **0.63** | 0.33 | 0.23 |
| 8) Caring | **0.67** | 0.28 | 0.24 |
| 9) Understanding | **0.68** | 0.30 | 0.20 |
| 10) Respect | **0.60** | 0.32 | 0.29 |
| *Instructions: Now you will hear different ways that women relate to one another. As you hear each word, please indicate if you experienced this with other women at ANC.* |  |  |  |
| 11) Companionship | **0.62** | 0.18 | 0.44 |
| 12) Belonging | **0.85** | -0.06 | 0.33 |
| 13) Shared activities | **0.98** | -0.11 | 0.17 |
| 14) Shared experience | **0.81** | 0.06 | 0.29 |
| *Instructions: The next set of words are about exchanging information. As I read each word, please tell me if you rechanged information in these ways* |  |  |  |
| 15) Exchanged advice | **0.89** | 0.003 | 0.20 |
| 16) Exchanged guidance/suggestions | **0.88** | -0.01 | 0.23 |
| 17) Exchanged useful information | **0.98** | -0.13 | 0.18 |

#### Provider Connectedness

The PRES was developed by a research team in the US to describe how women in group antenatal care experience empowerment; it includes four theoretical domains, one of which is provider connectedness. The scale was validated in English and in Spanish [35]. The provider connectedness subscale includes questions about being able to ask the midwife questions and being listened to and respected; thus, all women receiving antenatal care may report feeling connected to their provider. The PRES has been translated to Chichewa and previously used in Malawi [36]. The individual items are in Table 4.

**Table 4:** The items of the provider connectedness scale and their CFA loading and uniqueness.

|  | Factor 1 | Uniqueness |
|---|---|---|
| *Instructions: The following questions are in reference to your antenatal care and interactions with midwives at antenatal care. Please listen to each statement carefully and then choose the response that best describes how strongly your agree or disagree with the statement.* |  |  |
| 1) I can ask my midwife provider about my pregnancy. | **0.55** | 0.70 |
| 2) I have enough time with my midwife to discuss my pregnancy. | **0.74** | 0.45 |
| 3) My midwife listens to me. | **0.80** | 0.36 |
| 4) My midwife respects me. | **0.82** | 0.32 |
| 5) I expect my midwife to respect my decisions about my pregnancy. | **0.74** | 0.46 |
| 6) My midwife respects my decision, even if it is different than her/his recommendation. | **0.57** | 0.67 |

**Table 5:** Results of the GLM in transformed scale.

|  | Perinatal Peer Support Scale |  |  |  | Pregnancy Related Empowerment Scale |  |
|---|---|---|---|---|---|---|
|  | Relational Support |  | Peer Network |  | Provider connectedness |  |
|  | Exponentiated coefficients (SE) | *p* value | Exponentiated coefficients (SE) | *p* value | Exponentiated coefficients (robust SE) | *p* value |
| Assignment | 0.57 (0.03) | <0.001 | 0.83 (0.02) | <0.001 | 0.72 (0.03) | <0.001 |
| Parity (ref: primip) | 1.05 (0.08) | 0.526 | 1.02 (0.04) | 0.524 | 1.00 (0.05) | 0.955 |
| Age (ref: 15-19) |  |  |  |  |  |  |
| 20-24 | 0.96 (0.08) | 0.677 | 0.96 (0.04) | 0.283 | 0.94 (0.06) | 0.319 |
| 25-29 | 0.93 (0.10) | 0.520 | 0.95 (0.04) | 0.319 | 0.96 (0.08) | 0.621 |
| 30-34 | 0.88 (0.11) | 0.286 | 0.95 (0.05) | 0.342 | 0.85 (0.08) | 0.074 |
| 35 + | 0.94 (0.13) | 0.644 | 0.92 (0.06) | 0.200 | 1.09 (0.11) | 0.410 |
| **Clinic (ref: 0)** |  |  |  |  |  |  |
| 1 | 1.08 (0.11) | 0.424 | 1.02 (0.05) | 0.702 | 1.13 (0.09) | 0.147 |
| 2 | 1.15 (0.11) | 0.130 | 1.00 (0.05) | 0.980 | 1.08 (0.08) | 0.280 |
| 3 | 1.27 (0.13) | 0.017 | 1.01 (0.05) | 0.822 | 1.12 (0.09) | 0.154 |
| 4 | 1.34 (0.14) | 0.005 | 1.08 (0.05) | 0.107 | 1.09 (0.09) | 0.310 |
| 5 | 1.46 (0.14) | <0.001 | 1.13 (0.05) | 0.007 | 1.17 (0.09) | 0.048 |
| 6 | 1.42 (0.16) | 0.002 | 1.15 (0.06) | 0.011 | 1.47 (0.13) | <0.001 |
| Education (ref: less than primary) |  |  |  |  |  |  |
| Completed primary | 0.67 (0.04) | <0.001 | 0.88 (0.03) | < 0.001 | 1.04 (0.04) | 0.397 |
| More than primary | 0.67 (0.06) | <0.001 | 0.86 (0.04) | 0.003 | 1.11 (0.08) | 0.148 |

**Outcomes**: The outcome measures (relational support, peer network, and provider connectedness) were reverse-transformed (max + 1 – original score). Lower transformed values indicate **higher outcomes**.
**Model**: Gamma family with log link for relational support and provider connectedness. Gaussian family with log link for peer network. Results are exponentiated coefficients (exp(β)), interpreted as multiplicative changes in the transformed outcome

#### Health Outcomes

Clinical outcomes on the impact of group antenatal care compared to individual antenatal care are birth weight and maternal psychological distress, measured at T3 (2 month postpartum). The original outcome of the trial was preterm birth, but challenges in accurately measuring gestational age led to the use of birth weight as a proxy for preterm birth. Birth weight was measured at birth on a scale at the health facility and then collected from health records by research assistants.

Psychological distress was measured by the Self-Reporting Questionnaire (SRQ-20), which is comprised of 20 yes/no questions [37]. An example item from the SRQ-20 is “Do you feel nervous, tense, or worried?” This instrument was validated in Malawi in the Chichewa language [38].

The following covariates were included in the model: clinic, age groups, education category, parity, relationship status, wealth quantile, and the ownership index (yes/no for 3 questions). Parity refers to birth history, with primiparous meaning of first birth and multiparous for subsequent births. The wealth quantile and ownership index were used as proxies for socioeconomic status. These data were collected at baseline for each participant in both group and individual antenatal care groups.

### Sample

One thousand eight hundred eighty-seven (N = 1887) pregnant women (less than 24 weeks at first antenatal visit) between the ages of 15 and 42 years were recruited, consented, and enrolled in the parent study. As noted above, eligible women were enrolled at their first antenatal visit by trained research assistants (RAs) and then randomized to receive individual antenatal care (N = 941) or group antenatal care (N = 946). Data are accessible in a public repository, University of Michigan - Deep Blue Data, at https://doi.org/10.7302/a315-2417.

### Statistical Analyses

Analyses were conducted in Stata (version 18) [39].

#### Power

This study included community-building measures collected at the end of the intervention (T2) and maternal and infant health outcomes collected at two months postpartum (T3). Based on the available sample size at those time points (n = 1686 at T2 and n = 1653 at T3), power analyses were conducted to confirm that the study was adequately powered to detect small effect sizes. Power calculations were completed assuming a statistical power of 0.8 and an alpha level of 0.05. Given these parameters the study was adequately powered to detect small effect sizes (Cohen’s d ≥ .15).

#### Balance of Randomization

All baseline measures were tested using chi-square tests to see if the randomization led to equivalency between the individual and group care groups. Any variables to which balance was not achieved during randomization were included as a covariate in the main analyses.

#### Differences in Missingness & Loss to follow up

Differences between those with and without missing data were tested with chi-square tests. Variables on which they differ were included in the analyses to reduce bias related to missingness. Similarly, to identify differences in those lost to follow up, baseline demographic variables were tested using t-tests and chi-squared tests among those lost between the start of the intervention (T1), the end of intervention (T2) and 2 months postpartum (T3) compared to those who remained in the study. The approach to handling missing data was determined after reviewing the distribution of missing data. Given that 20% of data were missing for key variables, multiple imputation by chained equations was used after testing that the data satisfied the missing at random assumption, which is appropriate for longitudinal data with both continuous and binary variables [40]. Sensitivity analyses were completed to ensure imputed variables were plausible. Variables that were different due to missingness or loss to follow up were included in analysis as covariates.

### Factor analysis (Aim 1a)

Exploratory factor analysis with tetrachoric correlations and principal component analysis was conducted to determine the construct underlying factor structure for the perinatal peer support scale [41]. The tetrachoric correlation matrix was computed to estimate the underlying continuous correlation between binary items, which is necessary because using Pearson correlations with binary data is inappropriate. The resulting matrix is then used to perform exploratory factor analysis to identify latent factors, similar to how continuous data is analyzed [42–44]. To determine the number of factors, we examined the number of Eigenvalues greater than 1 and a scree plot compared to randomized values from a parallel analysis, as well as the cumulative percentage of variability explained [45].

The provider connectedness subscale of the PRES was previously validated and utilized in Malawi, so confirmatory factor analysis was conducted with a one factor structure [35,36,46]. The threshold for factor loadings are above 0.4 indicating acceptable item performance. Model fit was assessed using multiple indices, including the comparative fit index (CFI) and Tucker–Lewis index (TLI), with values ≥0.90 indicating acceptable fit, and the root mean square error of approximation (RMSEA) and standardized root mean square residual (SRMR), with values ≤0.08 indicating acceptable fit [47,48].

To develop a comprehensive community building measure, higher order factor analysis was attempted to test the validity of a higher-order multidimensional community building construct comprised of the two scales (perinatal peer support and provider connectedness) [49]. The outcomes of this analysis determined the final factor structure of community building used to answer both research aims comparing the community building scores of the group and individual antenatal care groups and examining the effect of community building on maternal and infant outcomes through mediation analyses.

### Main Regression Analysis (Aim 1b)

Before conducting the main analyses, the distribution of the variables was examined for outliers and to determine whether they meet the assumptions of planned analyses. If they did not meet the assumptions, generalized linear models allowing for alternative distributions (Poisson, Gamma, Negative Binomial) were considered for the main analyses. Analyses were conducted in Stata (version 18) [39].

#### Regression Analysis

One subscale of the perinatal peer support scale (relational support) and the provider connectedness measure were left-skewed, violating the distribution assumptions for regression models. Therefore, the outcomes were reverse transformed (*Transformed Y* = (max(*Y*) +1) ― (*Y*)). This positive, right-skewed data was appropriate with a gamma distribution with a log link. The other subscale of the perinatal peer support scale (network) was evenly distributed. To maintain consistency for the interpretation, this was reversed transformed (*Transformed Y* = (max(*Y*) +1) ― (*Y*)), similar to the other perinatal peer support subscale. Because of this normal distribution, a Gaussian GLM was used with a log link. Covariates were included where significant and based on the results of missingness and loss to follow up analysis.

### Mediation Analysis (Aim 2)

The primary outcomes of this analysis were psychological distress (SRQ-20) and birth weight, collected at two months postpartum (T3), mediated by community building measures of both perinatal peer support subscales (relational support and peer network) and the provider connectedness subscale. The independent variable was intervention status. Generalized structural equation modeling was used to test the model with all the mediators and then separately including covariates [50].

Nonlinear combination of parameters was used as a post-estimation command to test the direct and indirect effects of the mediation model. Covariates included variables shown in previous research to be related to psychological distress or pre-term birth (age, marital status, education, parity, socioeconomic status) as well as other baseline variables related to imbalance in randomization, loss to follow up, or differences in missingness.

## Results

The parent study consented and enrolled 1887 eligible pregnant women, with N = 946 randomized to group antenatal care intervention and N = 941 to individual antenatal care (see Table 2). Of the analytic sample, greater than 40% of women in both groups were primiparous and the vast majority (94%) were in a relationship. Approximately 45% reported less than a primary school education, while 40% had completed primary school. In the ownership index, which asked whether participants had electricity or a generator, farm animals or poultry, and agricultural land, 20% had none of these.

Chi-squared tests of each of the baseline demographic variables to test for balance in randomization indicated that there were no significant differences between the group and individual antenatal care groups (see Table 2).

At baseline, 1887 participants were enrolled in the trial. At T2 (end of intervention), 1686 participants were in the trial, and 198 participants were lost to follow up. At T3 (2 months postpartum), 1653 participants were in the trial, and 231 were lost to follow up. Using chi-square analyses, those in the trial and the lost to follow up were different from baseline in assignment (*p* = 0.005), parity (*p* < 0.001), age (*p* < 0.001), and clinic (*p* < 0.001).

The majority of the 1686 women in the group and individual antenatal care groups at T2, 1,402 (83%) completed the perinatal peer support and provider connectedness questions. There are significant differences in those who completed both the perinatal peer support and provider connectedness questions on age (*p* = 0.001), clinic, (*p* < 0.001), and parity (*p* < 0.001). After multiple imputation by chained equations was used, sensitivity analyses were completed which ensured imputed variables were plausible.

### Results for Aim 1a: Factor analysis

#### Perinatal peer support

(Aim 1a): A two factor model was selected for the exploratory factor analysis based on the number eigenvalues greater than 1, scree plot, and the cumulative percentage of variability explained. The eigenvalues are 10.83 for Factor 1 and 1.07 for Factor 2, with a cumulative variance of 83.8% explained by the two factors. The factor loadings were all above 0.54, and the uniqueness below 0.48 for all items. Table 3 shows each item with their loading and uniqueness. The two subscales were significantly and positively correlated (0.53).

The two factors were named 1. “Relational Support” and 2. “Peer Network.” The two subscales cover key aspects of community building: the relational support exploring the emotional support, relating among women, and the exchange of information and experiences that women experience during antenatal care; and the peer network subscale focusing on the friendships that women develop during antenatal care and how that extends beyond group.

The confirmatory factor analysis for the provider connectedness scale, showed a good fit for the model, and was confirmed as a one factor model with factor loadings ranging from 0.55-0.80, above the threshold of 0.4 and uniqueness between 0.32 and 0.7, demonstrating acceptable item performance. The comparative fit index (CFI = 0.95) and Tucker–Lewis index (TLI = 0.91) were above the conventional standard of ≥ 0.90 and the standardized root mean square residual (SRMR = 0.07) was within the acceptable range, while the root mean square error of approximation (RMSEA = 0.12) exceeded the acceptable cutoff of 0.08 [47]. Given the strong factor loadings, internal consistency of 0.85 (Cronbach’s alpha), and other satisfactory fit indices, the model has adequate construct validity.

The model to create a comprehensive community building measure by combining the perinatal peer support subscale and provider connectedness using a higher order factor analysis, did not converge. Therefore, we proceeded using two sub-scales of perinatal peer support and the provider connectedness scale as separate variables in the regression and mediation models.

In summary, the results for Aim 1a, the factor analysis found adequate evidence for the construct validity of the perinatal peer support scale with two subscales and confirmed a one factor model for the provider connectedness. The comprehensive community building measure did not converge.

### Results for Aim 1b: Regression

Participants in group antenatal care had better (lower scores, given the transformation of the scales) scores for the two perinatal peer support subscales: relational support and peer network as well as provider connectedness than the individual antenatal care group. Participants in group antenatal care had significantly lower scores on the relational support subscale of the perinatal peer support scale, indicating more relational support, with a 43% decrease in the mean (p < 0.001) in relational support compared with those in individual antenatal care, controlling for covariates of parity, age, clinic, and education. We tested the assumptions for the Gamma GLM model with log link function using multiply imputed data. The model fit reasonably well, with mild overdispersion (Pearson χ²/df =1.04), a deviance/df ratio below 1 (0.65), and appropriate deviance (right skewed, fitting with expectations for Gamma model) and Pearson residuals (no pattern across the predictor).

Peer network subscale of the perinatal peer support scale was significantly lower for participants in group antenatal care than individual care, indicating stronger peer network, with a 17% lower mean in the transformed scale (p < 0.001) controlling for covariates of parity, age, clinic, and education. Since the peer network sub-scale was not as skewed, the Gaussian distribution was appropriate. The identity link function is most commonly used with a Gaussian distribution, however, for consistency in interpretation we decided to use the log link function, which is also acceptable since the outcome is strictly positive and model fits the data well. The Gaussian GLM with log link fit reasonably well, with mild overdispersion (Pearson χ²/df =1.14), a deviance/df ratio just above 1 (1.14), roughly normal deviance residuals and the Pearson residuals showing no pattern across the predictor.

The scores for provider connectedness were significantly lower for those in the group antenatal care than individual antenatal care, indicating stronger connection between provider and patient, with a 28% (p < 0.001) difference in the transformed scale controlling for covariates of parity, age, clinic, and education. The Gamma GLM with log link fit acceptably, showing signs of under dispersion (Pearson χ²/df =0.68) and a deviance/df ratio below 1 (0.67). Deviance residuals were right-skewed, consistent with the expectations for a Gamma model, and Pearson residuals showed no pattern across the predictor indicating that the distribution was appropriate.

### Results for Aim 2: Mediation Analysis

In the generalized structural equation model (GSEM) with the outcome of psychological distress, mediated by community building (subscales of perinatal peer support scale and the provider connectedness scale) using multiply imputed data, neither the direct relationship (*β* = 0.98, *p* = 0.906) between intervention and psychological distress scores nor the indirect relationships mediated by community building (relational support, *β* = 1.00, *p* = 0.956; peer network, *β* = 0.99, *p* = 0.167; and provider connectedness, *β* = 1.00, *p* = 0.494) were significant. Furthermore, none of the direct associations between the community building and the outcomes of psychological distress were significant (relational support, *β* = 1.00, *p* = 0.817; peer network, *β* = 1.03, *p* = 0.159; and provider connectedness, *β* = 0.99, *p* = 0.404).

Similarly, in the GSEM for the outcome of birthweight, mediated by community building, the direct relationship between group assignment and birthweight was not significant (*β* = 0.99, *p* = 0.119). The indirect relationship between group assignment mediated by community building was not significant (relational support, *β* = 1.00, *p* = 0.906; peer network, *β* = 0.99, *p* = 0.141; and provider connectedness, *β* = 1.00, *p* = 0.542). The relationship between community building and birthweight was also not significant (relational support, *β* = 1.00, *p* = 0.976; peer network, *β* = 0.99, *p* = 0.191; and provider connectedness, *β* = 1.00, *p* = 0.905).

## Discussion

Group antenatal care is an evidence-based model that has shown positive effects during the perinatal period, yet one of its core components, community building, is underexplored. This study begins to fill this gap by validating measures of perinatal peer support and provider connectedness and using them to assess community building within a randomized controlled trial of group antenatal care in Malawi. The CenteringPregnancy-based model was adapted and implemented across seven clinics in Blantyre District, where 1,887 participants were randomized to either group or individual antenatal care. After validating each measure, we found that women who participated in group antenatal care reported significantly stronger community building across all indicators. However, in the generalized structural equation model examining these outcomes, community building did not mediate the effect of group antenatal care on maternal psychological distress or infant birthweight. Together, these findings strengthen our ability to begin to measure aspects of community building and show that group antenatal care may meaningfully enhance community building among participants.

In this study, community building was measured with perinatal peer support, comprising two domains of relational support and peer network, as well as provider connectedness. Conceptually, these measures capture the peer and provider dimensions of community building. Both measures were left-skewed with limited variance, suggesting a ceiling effect in community building. The attempt to combine the measures into one overall community-building score through higher order factor analysis was not successful as they did not converge. This suggests the two measures capture different, though related, parts of community building. While this study examined the construct validity using exploratory factor analysis, it did not address other aspects of validity. In particular, future studies could address criterion validity using social support or other proximal health outcomes [51].

The perinatal peer support scale was developed by a research team as part of the preceding R21 pilot study; therefore, evidence regarding its use and psychosmetric performance remains limited to this program of research [22,52]. In contrast, the PRES, including the provider connectedness subscale, has been adapted and evaluated in studies in Turkey and Indonesia [53,54]. Both studies demonstrated acceptable construct validity, with the Turkish adaptation reporting higher factor loadings and fit statistics [53,54]. These findings support the continued use of the instrument to measure women’s perceptions of provider connectedness during pregnancy. This study focuses on community building within group antenatal care, using measures that assess the experiences of relational connections and support within group antenatal care.

Social support has also been used in the group antenatal care literature, with mixed findings: with some evidence of increased reliance on peer networks and improved psychological quality of life, but limited evidence that general social support mediates health outcomes [21,55,56]. There is an important difference between what these instruments are measuring, as these community building measure focus specifically on the experience of connecting to peers and providers during antenatal care, whereas social support is a perceived measure of support generally.

This study found a significant relationship between participation in group antenatal care and perinatal peer support (relational support and peer network), as well as provider connectedness. These findings align with the theoretical premise that group antenatal care fosters social connections and trust among participants as well as between women and providers. Grenier and colleagues’ theory of change identifies community building as one component of a broader transformation in providing group antenatal care. Together with increased time, decreased hierarchy, interactive learning, self-assessment, and continuity of care, community building is expected to lead to supportive relationships, better communication, empowered partnerships, and more meaningful care. These transformed experiences may contribute to a self-reinforcing cycle of more and better care, and through improved health behaviours, communication, and service use, create opportunities for improved maternal and newborn outcomes [32]. The peer support and provider connectedness measures examined here capture dimensions of this theory of change, but not the full mechanisms outlined. Accordingly, the mediation analysis tests one specific pathway, not the full theory of change.

However, the findings on community building remain mixed. In pilot RCTs in Malawi and Tanzania, significantly higher community building was found using measures of provider connectedness and peer connectedness, while a quasi-experimental study in Tanzania did not find significant differences between those in group and individual antenatal care on community building [36,57]. The literature in group antenatal care has mixed findings on social support outcomes, with one study in the US finding that group antenatal care participants were significantly more likely to depend on friendship or peer networks for social support (OR 2.81), while two studies did not find significant differences in social support between those in group and individual care [21,55,56].

In the present analyses, these specific measures of community building did not mediate the relationship between group antenatal care participation and maternal psychological distress or infant birthweight. The study demonstrating higher social support among those receiving group care also found that the social support mediated the relationship between group antenatal care and psychological quality of life, while another study found that social support did not mediate the relationship between care and postpartum depression by intervention group [55,58]. A future area of research could explore the pathway from community building to social support to health outcomes or through other more intermediate ways like knowledge change, engagement in care, or health behavior not included in the analyses.

This study adds to the growing literature on group antenatal care by establishing the validity of the Perinatal Peer Support Scale, with two distinct domains of relational support and peer network, and the Provider Connectedness scale within the Malawian context. The findings demonstrate significant improvements in these community building measures among women who participated in group antenatal care compared with those in individual care. Although the mediation analysis did not yield significant indirect effects on maternal psychological distress or infant birthweight, these results underscore the importance of continuing to develop and test theoretical models that link the core components of community building, such as trust, engagement, and social connectedness, to maternal and infant health outcomes as well as more proximal outcomes. Building and refining such models will be essential for understanding how community building in group antenatal care leads to changes in outcomes, through the strengthening of social relationships and collective efficacy among women, providers, and communities.

### Implications

#### Research Implications

This study contributes to the growing evidence that community building in group antenatal care can be measured and is strengthened through participation in group antenatal care. Validating the perinatal peer support and provider connectedness scales in the Malawian context provides tools to begin to quantify constructs central to community building. Further work is needed to refine mediation models and develop a pathway linking community building to health outcomes through direct and indirect pathways. Future research should test longitudinal models that explore community building’s relationship to proximal and distal health outcomes.

#### Practice Implications

For practitioners of group antenatal care, the results highlight the value of intentionally designing group antenatal care sessions for community building, fostering bonding among women and linking relationships with providers. Training for midwives and co-facilitators should emphasize facilitation skills that strengthen community building, and the use of relational care may be an approach that would further advance group antenatal care. Relational care is an approach to care that prioritizes the building of meaningful relationships based on respect, trust, and inclusivity, recognizing that human connection is essential for wellbeing. Unlike traditional person-centered care, which can be seen as a collection of needs, relational care focuses on the quality of interactions between the person receiving care, the caregivers, their families, and the community. The goal of relational care is to foster a sense of autonomy, security, and self-worth for everyone involved in the care process.

#### Policy Implications

The findings suggest that the consequences stemming from community building in group antenatal care can meaningfully enhance women’s sense of support, network, and connectedness with peers and providers. As group antenatal care expands, policy-level efforts are needed to preserve community building as a core component to ensure high-quality implementation. Furthermore, investing in the expansion of group antenatal care through insurance coverage and implementation support globally, while maintaining the core components of group antenatal care, may expand access to high-quality antenatal care and could improve mental and physical health during the perinatal period.

### Limitations

This analysis was based on secondary data collected as part of a randomized controlled trial, which created limitations as the measures were constrained by the parent study design, available variables and measures, and timing of data collection. Some constructs relevant to community building, such as the quality of group facilitation, the depth of peer interaction, or longitudinal changes in peer or provider relationships, were not captured.

Additionally, the provider connectedness subscale was not originally developed for the Malawian context, which may have influenced item interpretation despite demonstrated internal consistency and factor validity. Both community building scales were significantly left-skewed. Although the generalized linear models accounted for these skewed distributions, there may have been ceiling effects, a measurement limitation where many participants clustered at the highest possible score on community building making it difficult to measure true differences. In addition, the RMSEA for the provider connectedness model exceeded the conventional cutoff, although the other fit indices were acceptable. Finally, causality cannot be inferred beyond the scope of the original trial design, and residual confounding may remain despite statistical adjustment.

## Data Availability

Data are accessible in a public repository, University of Michigan - Deep Blue Data, at https://doi.org/10.7302/a315-2417

## Acknowledgements

The authors thank the women who participated in this study and the midwives, community facilitators, and research assistants at the seven participating clinics in Blantyre District. AB is grateful to the PhD Program at the Johns Hopkins School of Nursing, especially Program Director Dr. Kamila Alexander; to Dr. Sarah Murray for her support and for serving as chair of her dissertation committee; and to Dr. Melissa Hladek, whose thoughtful guidance helped shape this study.

---

**S1 Table:** CONSORT 2025 checklist of information to include when reporting a randomised trial.

| Section / Topic | No | CONSORT 2025 checklist item description | Reported on page no. |
|---|---|---|---|
| **Title and abstract** |  |  |  |
| Title and structured abstract | 1a | Identification as a randomised trial | 1 |
|  | 1b | Structured summary of the trial design, methods, results, and conclusions | 2 |
| **Open science** |  |  |  |
| Trial registration | 2 | Name of trial registry, identifying number (with URL) and date of registration | 7 |
| Protocol and statistical analysis plan | 3 | Where the trial protocol and statistical analysis plan can be accessed | 7 |
| Data sharing | 4 | Where and how the individual de-identified participant data (including data dictionary), statistical code and any other materials can be accessed | 12 |
| Funding and conflicts of interest | 5a | Sources of funding and other support (e.g., supply of drugs), and role of funders in the design, conduct, analysis and reporting of the trial | Reporting in system |
|  | 5b | Financial and other conflicts of interest of the manuscript authors | Reporting in system |
| **Introduction** |  |  |  |
| Background and rationale | 6 | Scientific background and rationale | 4-7 |
| Objectives | 7 | Specific objectives related to benefits and harms | 7 |
| **Methods** |  |  |  |
| Patient and public involvement | 8 | Details of patient or public involvement in the design, conduct and reporting of the trial | n/a |
| Trial design | 9 | Description of trial design including type of trial (e.g., parallel group, crossover), allocation ratio, and framework (e.g., superiority, equivalence, non-inferiority, exploratory) | 7 |
| Changes to trial protocol | 10 | Important changes to the trial after it commenced including any outcomes or analyses that were not prespecified, with reason | n/a |
| Trial setting | 11 | Settings (e.g., community, hospital) and locations (e.g., countries, sites) where the trial was conducted | 7 |
| Eligibility criteria | 12a | Eligibility criteria for participants | 8 |
|  | 12b | If applicable, eligibility criteria for sites and for individuals delivering the interventions (e.g., surgeons, physiotherapists) | n/a |
| Intervention and comparator | 13 | Intervention and comparator with sufficient details to allow replication. If relevant, where additional materials describing the intervention and comparator (e.g., intervention manual) can be accessed | n/a |
| Outcomes | 14 | Pre-specified primary and secondary outcomes, including the specific measurement variable (e.g., systolic blood pressure), analysis metric (e.g., change from baseline, final value, time to event), method of aggregation (e.g., median, proportion), and time point for each outcome | 8-12 |
| Harms | 15 | How harms were defined and assessed (e.g., systematically, non-systematically) | n/a |
| Sample size | 16a | How sample size was determined, including all assumptions supporting the sample size calculation | 12 |
|  | 16b | Explanation of any interim analyses and stopping guidelines | n/a |
| Randomisation: |  |  |  |
| Sequence generation | 17a | Who generated the random allocation sequence and the method used | n/a |
|  | 17b | Type of randomisation and details of any restriction (e.g., stratification, blocking and block size) | n/a |
| Allocation concealment mechanism | 18 | Mechanism used to implement the random allocation sequence (e.g., central computer/telephone; sequentially numbered, opaque, sealed containers), describing any steps to conceal the sequence until interventions were assigned | n/a |
| Implementation | 19 | Whether the personnel who enrolled and those who assigned participants to the interventions had access to the random allocation sequence | n/a |
| Blinding | 20a | Who was blinded after assignment to interventions (e.g., participants, care providers, outcome assessors, data analysts) | n/a |
|  | 20b | If blinded, how blinding was achieved and description of the similarity of interventions | n/a |
| Statistical methods | 21a | Statistical methods used to compare groups for primary and secondary outcomes, including harms | 12-16 |
|  | 21b | Definition of who is included in each analysis (e.g., all randomised participants), and in which group | 16-17 |
|  | 21c | How missing data were handled in the analysis | 13 |
|  | 21d | Methods for any additional analyses (e.g., subgroup and sensitivity analyses), distinguishing prespecified from post-hoc | 13 |
| **Results** |  |  |  |
| Participant flow, including flow diagram | 22a | For each group, the numbers of participants who were randomly assigned, received intended intervention, and were analysed for the primary outcome | n/a – in original results |
|  | 22b | For each group, losses and exclusions after randomisation, together with reasons | n/a |
| Recruitment | 23a | Dates defining the periods of recruitment and follow-up for outcomes of benefits and harms | n/a – in original results |
|  | 23b | If relevant, why the trial ended or was stopped | n/a |
| Intervention and comparator delivery | 24a | Intervention and comparator as they were actually administered (e.g., where appropriate, who delivered the intervention/comparator, how participants adhered, whether they were delivered as intended [fidelity]) | n/a |
|  | 24b | Concomitant care received during<br>the trial for each group | n/a |
| Baseline data | 25 | A table showing baseline demographic and clinical characteristics for each group | 16 |
| Numbers analysed, outcomes and estimation | 26 | For each primary and secondary outcome, by group:<br>● the number of participants included in the analysis<br>● the number of participants with available data at the outcome time point<br>● result for each group, and the estimated effect size and its precision (such as 95% confidence interval)<br>● for binary outcomes, presentation of both absolute and relative effect size | 16 |
| Harms | 27 | All harms or unintended events in each group | n/a |
| Ancillary analyses | 28 | Any other analyses performed, including subgroup and sensitivity analyses, distinguishing pre-specified from post-hoc | 18 |
| **Discussion** |  |  |  |
| Interpretation | 29 | Interpretation consistent with results, balancing benefits and harms, and considering other relevant evidence | 26-30 |
| Limitations | 30 | Trial limitations, addressing sources of potential bias, imprecision, generalisability, and, if relevant, multiplicity of analyses | 31-32 |

**Supplemental Table 2:** Responses for the Perinatal Peer Support scale by group.

| Characteristic, n (%) | Individual antenatal care<br>n (%) | Group antenatal care<br>n (%) | *p*-value |
|---|---|---|---|
| 1) While at antenatal care, did you make new friends? | 423 (62.1) | 602 (83.3) | <0.001 |
| 2) Do you talk to any of these friends on the phone? | 256 (37.6) | 387 (53.5) | <0.001 |
| 3) Do you visit any of these friends? | 327 (48.0) | 411 (56.9) | 0.001 |
| *Instructions: As section will focus on relationships you developed with other women while attending clinic. First, you will hear a list of words that are about the kinds of emotional support that women sometimes get from one another. As you hear each word, please indicate if you experienced this type of support from other pregnant women at ANC.* |  |  |  |
| 4) Love/Affection | 595 (87.4) | 681 (94.2) | <0.001 |
| 5) Trust | 536 (78.7) | 653 (90.3) | <0.001 |
| 6) Acceptance | 503 (73.9) | 629 (87.0) | <0.001 |
| 7) Encouragement | 601 (88.3) | 685 (94.7) | <0.001 |
| 8) Caring | 567 (83.3) | 667 (92.3) | <0.001 |
| 9) Understanding | 578 (84.9) | 684 (94.6) | <0.001 |
| 10) Respect | 616 (90.5) | 698 (96.5) | <0.001 |
| *Instructions: Now you will hear different ways that women relate to one another. As you hear each word, please indicate if you experienced this with other women at ANC.* |  |  |  |
| 11) Companionship | 576 (84.6) | 675 (93.4) | <0.001 |
| 12) Belonging | 541 (79.4) | 683 (94.5) | <0.001 |
| 13) Shared activities | 533 (78.3) | 667 (92.3) | <0.001 |
| 14) Shared experience | 528 (77.8) | 673 (93.1) | <0.001 |
| *Instructions: The next set of words are about exchanging information. As I read each word, please tell me if you rechanged information in these ways* |  |  |  |
| 15) Exchanged advice | 590 (86.6) | 701 (97.0) | <0.001 |
| 16) Exchanged guidance/suggestions | 579 (85.0) | 693 (95.9) | <0.001 |
| 17) Exchanged useful information | 582 (85.5) | 691 (95.6) | <0.001 |

**Supplemental Table 3:** Responses for the provider connectedness scale.

| Characteristic, n (%) | Individual antenatal care<br>n (%) | Group antenatal care<br>n (%) | *p*-value |
|---|---|---|---|
| *Instructions: The following questions are in reference to your antenatal care and interactions with midwives at antenatal care. Please listen to each statement carefully and then choose the response that best describes how strongly your agree or disagree with the statement.* |  |  |  |
| 1) I can ask my midwife provider about my pregnancy. |  |  | 0.038 |
| Strongly Disagree | 6 (0.9) | 5 (0.7) |  |
| Disagree | 24 (3.5) | 11 (1.5) |  |
| Agree | 170 (25.0) | 159 (22.0) |  |
| Strong Agree | 481 (70.6) | 548 (75.8) |  |
| 2) I have enough time with my midwife to discuss my pregnancy. |  |  | <0.001 |
| Strongly Disagree | 9 (1.3) | 4 (0.6) |  |
| Disagree | 40 (5.9) | 16 (2.2) |  |
| Agree | 257 (37.7) | 193 (26.7) |  |
| Strong Agree | 375 (55.1) | 510 (70.5) |  |
| 3) My midwife listens to me. |  |  | <0.001 |
| Strongly Disagree | 8 (1.2) | 7 (1.0) |  |
| Disagree | 37 (5.4) | 10 (1.4) |  |
| Agree | 239 (35.1) | 175 (24.2) |  |
| Strong Agree | 397 (58.3) | 531 (73.4) |  |
| 4) My midwife respects me. |  |  | <0.001 |
| Strongly Disagree | 5 (0.7) | 6 (0.8) |  |
| Disagree | 35 (5.1) | 6 (0.8) |  |
| Agree | 238 (35.0) | 175 (24.2) |  |
| Strong Agree | 403 (59.2) | 535 (74.1) |  |
| 5) I expect my midwife to respect my decisions about my pregnancy. |  |  | <0.001 |
| Strongly Disagree | 9 (1.3) | 6 (0.8) |  |
| Disagree | 37 (5.4) | 8 (1.1) |  |
| Agree | 235 (34.5) | 187 (25.9) |  |
| Strong Agree | 400 (58.7) | 522 (72.2) |  |
| 6) My midwife respects my decision, even if it is different than her/his recommendation. |  |  | <0.001 |
| Strongly Disagree | 9 (1.3) | 5 (0.8) |  |
| Disagree | 37 (5.4) | 8 (1.1) |  |
| Agree | 235 (34.5) | 187 (25.9) |  |
| Strong Agree | 400 (58.7) | 522 (72.2) |  |

